# In-depth Analysis of Laboratory Parameters Reveals the Interplay Between Sex, Age and Systemic Inflammation in Individuals with COVID-19

**DOI:** 10.1101/2020.08.07.20170043

**Authors:** Felipe ten-Caten, Patrícia Gonzalez-Dias, Ícaro Castro, Rodrigo L T Ogava, Jeevan Giddaluru, Juan Carlo S Silva, Felipe Martins, André N A Gonçalves, André G Costa-Martins, José D. Araujo, Ana Carolina Viegas, Fernando Q Cunha, Sandra Farsky, Fernando A Bozza, Anna S Levin, Pia S Pannaraj, Thushan I de Silva, Paola Minoprio, Bruno B Andrade, Fabiano Pinheiro da Silva, Helder I Nakaya

## Abstract

**Introduction:** The progression and severity of the coronavirus disease 2019 (COVID-19), an infectious disease caused by the severe acute respiratory syndrome coronavirus 2 (SARS-CoV-2), varies significantly in the population. While the hallmarks of SARS-CoV-2 and severe COVID-19 within routine laboratory parameters are emerging, little is known about the impact of sex and age on these profiles.

**Methods:** We performed multidimensional analysis of millions of records of laboratory parameters and diagnostic tests for 178,887 individuals, of which 33,266 tested positive for SARS-CoV-2. These included complete blood cell count, electrolytes, metabolites, arterial blood gases, enzymes, hormones, cancer biomarkers, and others.

**Results:** COVID-19 induced similar alterations in the laboratory parameters in males compared to females. Biomarkers of inflammation, such as C-reactive protein (CRP) and ferritin, were increased especially in older men with COVID-19, whereas other markers such as abnormal liver function tests were common across several age groups, except for young women. Low peripheral blood basophils and eosinophils were also more common in the elderly with COVID-19. Both male and female COVID-19 patients admitted to the intensive care unit (ICU) displayed alterations in the coagulation system, and higher levels of neutrophils, CRP, lactate dehydrogenase (LDH), among others.

**Discussion:** Our study uncovers the laboratory profile of a large cohort of COVID-19 patients that underly discrepancies influenced by aging and biological sex. These profiles directly link COVID-19 disease presentation to an intricate interplay between sex, age and the immune response.

**One Sentence Summary:** Big Data analysis of laboratory results from a large number of COVID-19 patients and controls reveals distinct disease profiles influenced by age and sex which may underly occurrence of severe disease.

**Key messages:** *- What is the key question?:* Little is known about the impact of sex and age on the routine laboratory parameters measured in COVID-19 patients.

*- What is the bottom line?:* Our in-depth analysis unraveled distinct disease profiles influenced by age and sex which may underly occurrence of severe disease.

*- Why read on?:* This work will help physicians to interpret the disease presentation in COIVD-19 patients according to their age and sex.

## Introduction

The new coronavirus SARS-CoV-2 has rapidly spread throughout the world, causing an unprecedented pandemic. The virus is easily transmitted between humans and triggers a wide spectrum of clinical manifestations [1], ranging from asymptomatic or mild illness to severe disease (coronavirus disease 2019, COVID-19). SARS-CoV-2 infection has been associated with respiratory, gastrointestinal, hepatic and neurological dysfunction, which leads to acute respiratory distress syndrome, multiple organ failure, and death in severe cases [2].

Since SARS-CoV-2 is a systemic infection, inflammatory responses can induce changes in the cellular and biochemical composition of the peripheral blood [2]. For instance, a significant proportion of patients exhibit a variety of alterations in distinct organs, such as the gastrointestinal tract [3], liver [4], kidneys [5] and heart [6]. Such extra-pulmonary manifestations are associated with potential alterations in circulating levels of several biochemical parameters, such as bilirubin, urea, creatinine, myoglobin and coagulation factors.

To date, no systematic analysis of blood parameters has been performed on a large number of COVID-19 patients and particularly in the context of age and sex-related changes observed in the absence of SARS-CoV-2 infection. By analyzing millions of records of laboratory parameters from over 150,000 subjects, we revealed unique profiles of males and females with COVID-19 across different ages. Such large-scale analysis may provide critical information about COVID-19 pathogenesis related to sex and age. We also showed that the combination of several biochemical parameters and surrogates of inflammatory responses may be used to assess disease severity.

## Methods

### Dataset

We downloaded 3 datasets from the "COVID-19 Data Sharing/BR” repository which contained information about laboratory parameters and demographics, as well as diagnostic tests at https://repositoriodatasharingfapesp.uspdigital.usp.br/ (2020-06-30). The repository is an initiative of the São Paulo Research Foundation (FAPESP) in collaboration with the University of São Paulo and participation of Fleury Institute, Sírio-Libanês Hospital and Israelita Albert Einstein Hospital. We translated the names of the laboratory tests to English and identified common tests performed by the 3 sites. If a given test was performed more than once for the same subject, we retrieved the one collected at the earliest date after positive diagnosis of SARS-CoV-2 (by PCR or IgM). For the COVID-19 cases, we removed records collected before the day of positive diagnosis of SARS-CoV-2 or that were collected over 15 days the day of positive diagnosis of SARS-CoV-2. Laboratory parameters measured in less than 100 subjects or that contained discrete (i.e. categorical) values were removed from the analysis.

### Univariate analysis

We used the Anderson-Darling normality test to identify normally distributed laboratory parameters. The student t-test was used to identify the changes between group of samples, with p-values adjusted for multiple testing using the Benjamini & Hochberg method. The cutoffs utilized to identify a significant change were an Adjusted P-value < 0.05 and log2 fold-change > 0.4. Comparisons with less than 20 samples on both groups (i.e. COVID-19 cases and controls) were not considered. Plots were created with ggplot2 R package (https://ggplot2.tidyverse.org) and edited in CorelDraw software.

### Temporal age profiling

For each year of age (stratified by sex and COVID-19 diagnostic), we calculated the median and 95% confidence interval of each laboratory parameter. Using ggplot2 R package, we fitted a smooth line (method = loess) on the points and compared the lines between recent infection vs no-infection for each sex using Kolmogorov-smirnov test (FDR cutoff < 0.01). Ages with 2 or fewer values were not included in the median calculation.

### Intensive Care Unit (ICU) analysis

The Sírio-Libanês Hospital had identified data from patients admitted to the ICU (n= 285). For each subject, the first date labeled as when laboratory data were collected in the ICU was considered as day 0. We calculated the median and 95% confidence interval of each laboratory parameter on each day for male and female patients. Reference normal values of the parameters were obtained from physicians and displayed in the graphs. We grouped specific laboratory parameters into biomarkers of disease activity.

## Results

Laboratory parameter data from two hospitals and one diagnostics company in Brazil were made publicly available by the São Paulo Research Foundation (FAPESP). The repository holds open-access anonymized data from 175,887 individuals tested for SARS-CoV-2, of which 39,391 had tested positive using a combination of IgM/IgA/IgG serology and RT-PCR tests (Figure S1A). RT-PCR or IgM tests were positive in 33,266 subjects, suggesting recent infection with SARS-CoV-2, and were herein defined as COVID-19 cases. All remaining individuals were classified as controls, including the 6,125 subjects that have IgA (N = 900) or IgG (N=5,225) seroreactivity towards SARS-CoV-2 antigens. The age distributions of male and female COVID-19 cases were similar (Figure S1B). In total, we analyzed over 4.5 million records of 434 laboratory parameters and diagnostic tests (see methods).

**Figure S1.**
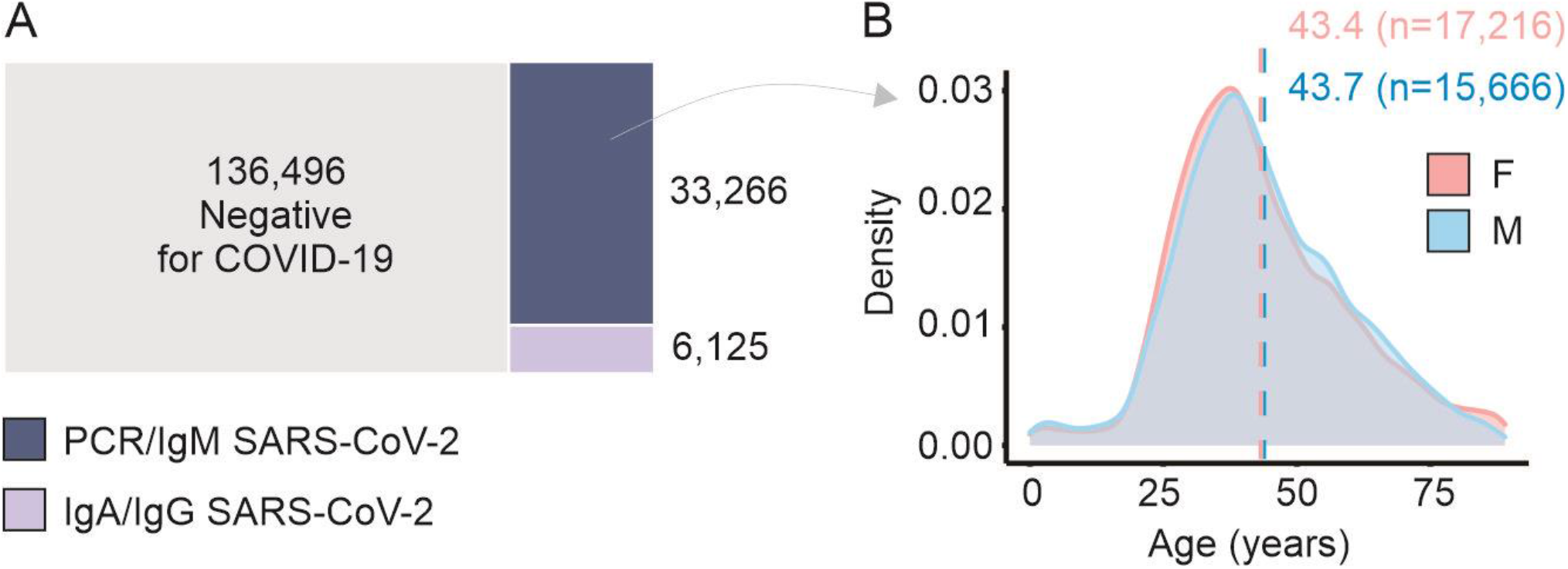
Laboratory parameters of individuals with COVID-19. **A**. Number of individuals with positive or negative diagnostic test result for SARS-CoV-2. The sizes of the rectangles are proportional to the number of individuals that were tested negative (grey) or positive (dark and light purple) for the virus. Subjects with positive tests for either PCR or IgM were considered as COVID-19 cases. **B**. Age distribution of COVID-19 cases. Dashed lines show the mean age in years of female (pink) and male (blue) subjects. No significant difference between both distributions was found by Kolmogorov-Smirnov test.

We divided the subjects into 3 age groups: 0 to 12 years old (N = 5,218), 13 to 60 years old (N = 146,404), and ≥61 years old (N = 22,657). Age information was not available for 1,608 individuals. We then performed univariate analyses comparing values of laboratory parameters between COVID-19 cases and controls within each age group. No significant differences were found between females and males in the 0 to 12 years group (Table S1). COVID-19 induced similar alterations in the laboratory parameters in males compared to females between 13 and 60 years old, in contrast to older individuals, where several parameters were altered by COVID-19 in both men and women (Figure 1). Male and female COVID-19 patients had a lower level of basophils and eosinophils, as well as significantly higher levels of gamma-glutamyl transferase (GGT), C-reactive protein (CRP) and ferritin when compared with control individuals in the same age group (Figure 1, Table S1).

**Figure 1.**
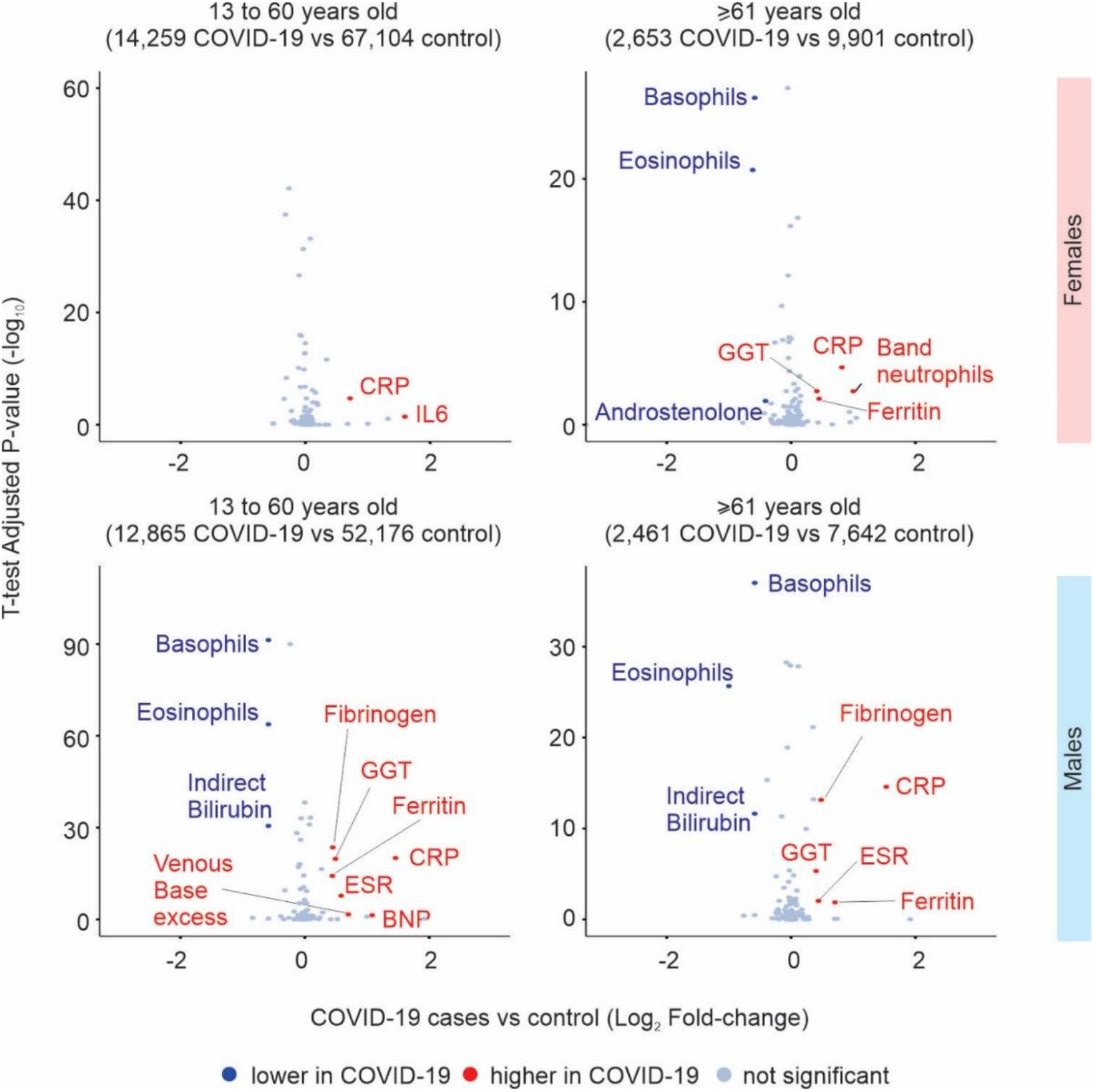
Alterations induced by COVID-19. Volcano plots showing the differences in the levels of laboratory parameters between COVID-19 cases and controls. Female subjects are shown on the top and male subjects on the bottom. The age group is indicated above each volcano plot and the number of individuals is shown in parenthesis. The x-axis shows the log2 fold-change between COVID-19 cases and controls, and the y-axis shows the −log10 adjusted P-value of the analyzed laboratory parameters (dots). Red and blue dots show, respectively, the laboratory parameters that were significantly higher or lower in COVID-19 cases compared to controls (Adjusted P-value < 0.05 and log2 fold-change > 0.4).

We generated a profile of key clinical chemistry parameters for male and female COVID-19 cases compared with control cases across age (Figure 2). Our analysis revealed important differences related to age and sex even in the absence of COVID-19. While alterations in several parameters were seen in COVID-19 cases, the absolute levels were critically dependent on whether patients were male or female, with the most perturbed laboratory markers seen in male COVID-19 cases (Figure 2).

**Figure 2.**
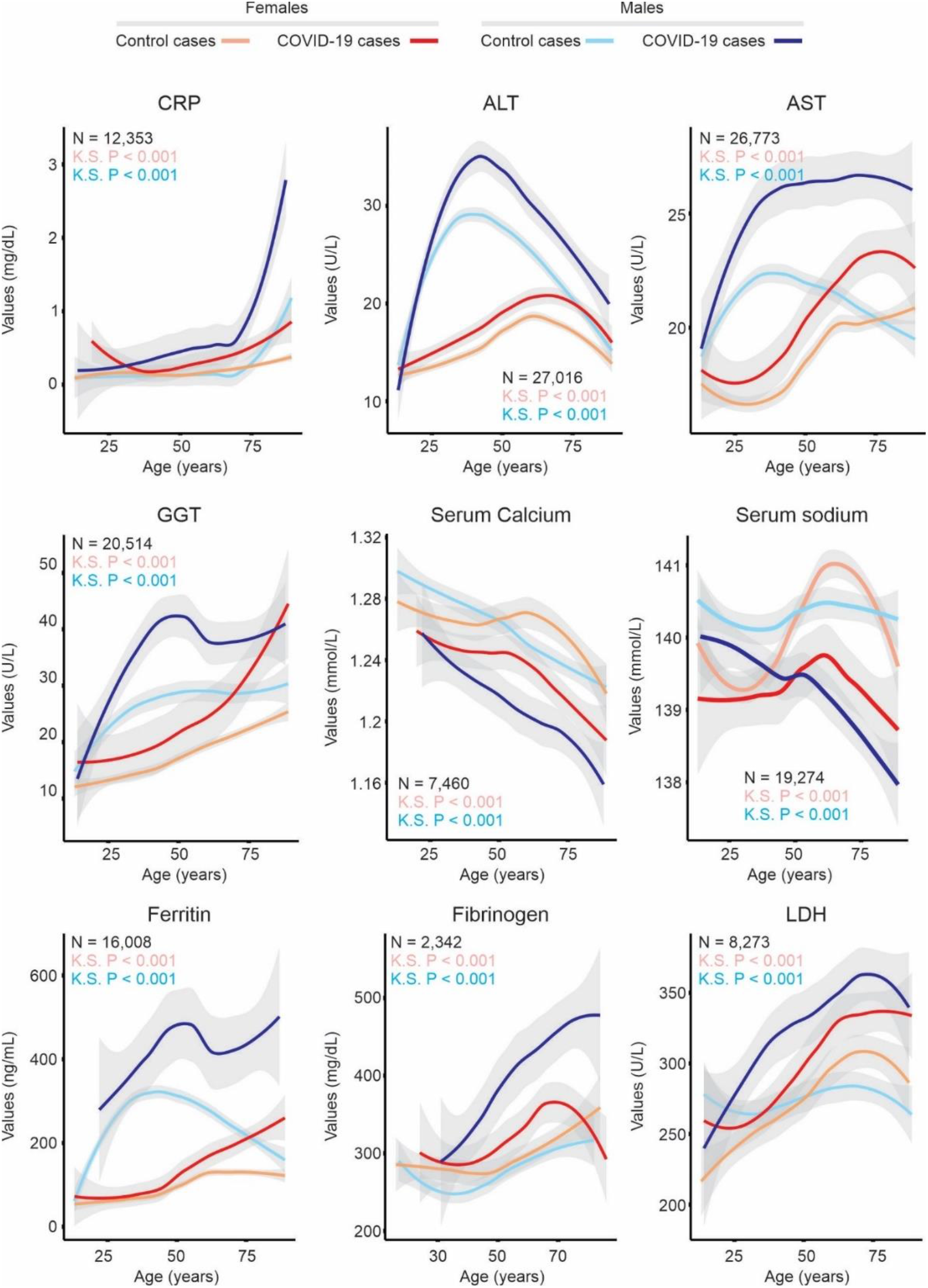
Selected laboratory parameter temporal profile of male and female COVID-19 patients. The x-axis shows the age of individuals and the y-axis shows the fitted median values (thick line) of the laboratory parameter displayed above each graph. The shaded area represents the 95% confidence interval. Female COVID-19 cases and controls are shown as red and brown lines, respectively. Male COVID-19 cases and control cases are shown as dark blue and light blue lines, respectively. The number of individuals (N) utilized in the analysis, and the Kolmogorov-Smirnov test p-value (K.S.) between the COVID-19 curve and the control curve for each sex are indicated.

We compared the absolute counts of cell populations in the complete blood count profile in COVID-19 cases and controls by sex and age (Figure 3). Lower counts of platelets, basophils, lymphocytes, and eosinophils were observed in COVID-19 cases compared to controls in both males and females (Figure 3).

**Figure 3.**
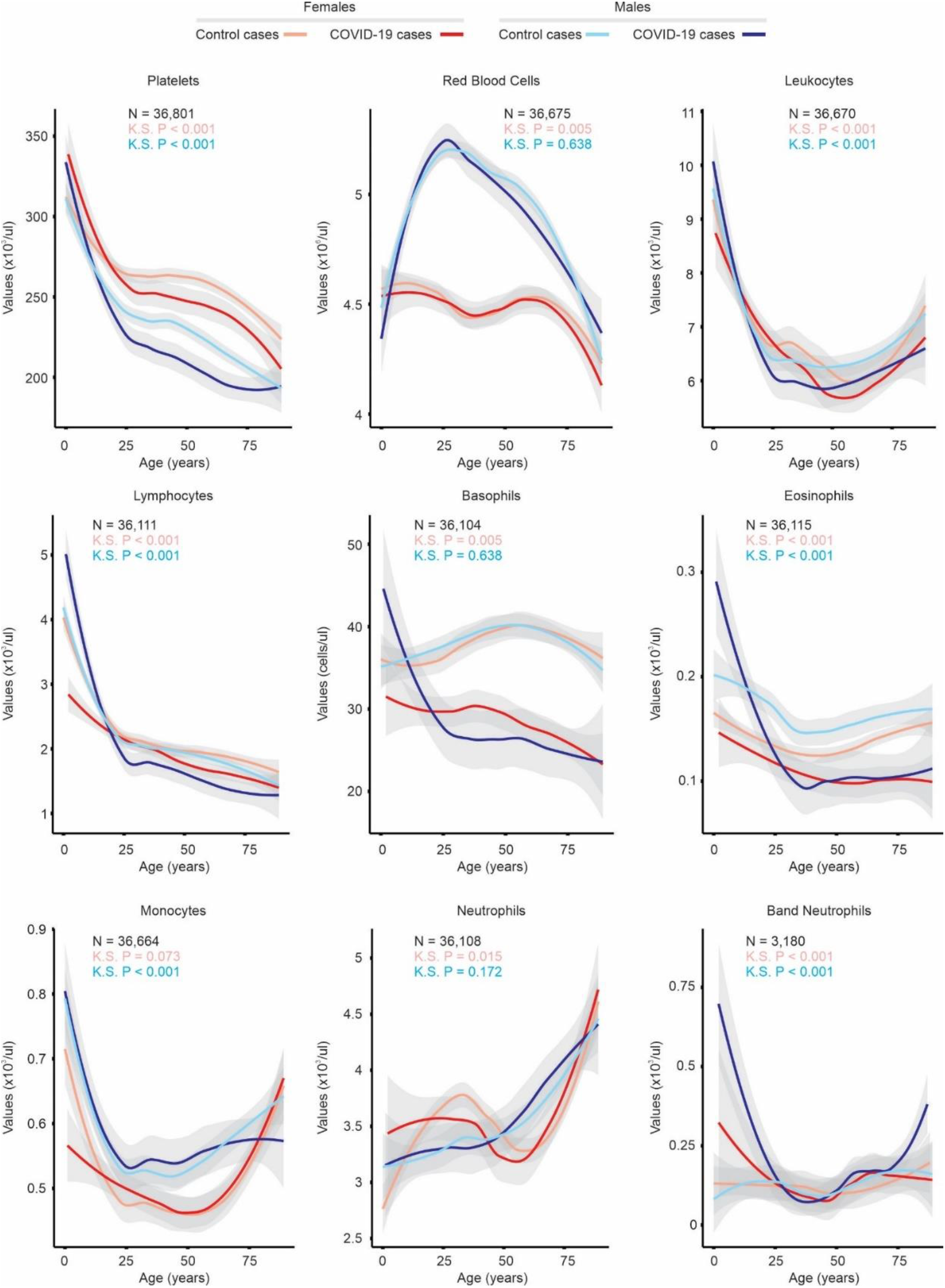
Analysis of complete blood count tests in COVID-19 cases. The x-axis shows the age of individuals and the y-axis shows the fitted median values (thick line) of the cell type displayed above each graph. The shade area represents the 95% confidence interval. Female COVID-19 cases and controls are shown as red and brown lines, respectively. Male COVID-19 cases and control cases are shown as dark blue and light blue lines, respectively.

The number of individuals (N) utilized in the analysis, and the Kolmogorov-Smirnov test p-value (K.S.) between the COVID-19 curve and the control curve for each sex are indicated.

Two hundred and eighty-five COVID-19 patients went to the intensive care unit (ICU). The median number of days in ICU was 15 (Figure 4A), and 74% of patients were male (Figure 4B), with 165 individuals aged ≥60 years of age (range 20 to 88) (Figure 4C). Daily changes in laboratory parameters during ICU stay were examined, with several remaining significantly altered throughout this period compared to reference values for age (Figure 4D and S2). Significant elevation of coagulation markers activated partial thromboplastin time (aPTT) and prothrombin time (PT/INR) was seen. While aPTT values detected in critically ill male COVID-19 patients returned back to normal after 30 days in ICU, those remained altered in female COVID-19 patients in ICU (Figure 4D). The curve of aPTT followed a similar trend as the curve for Lactate Dehydrogenase (LDH) (Figure 4D), reinforcing the link between activation of coagulation and of inflammatory pathways.

**Figure 4.**
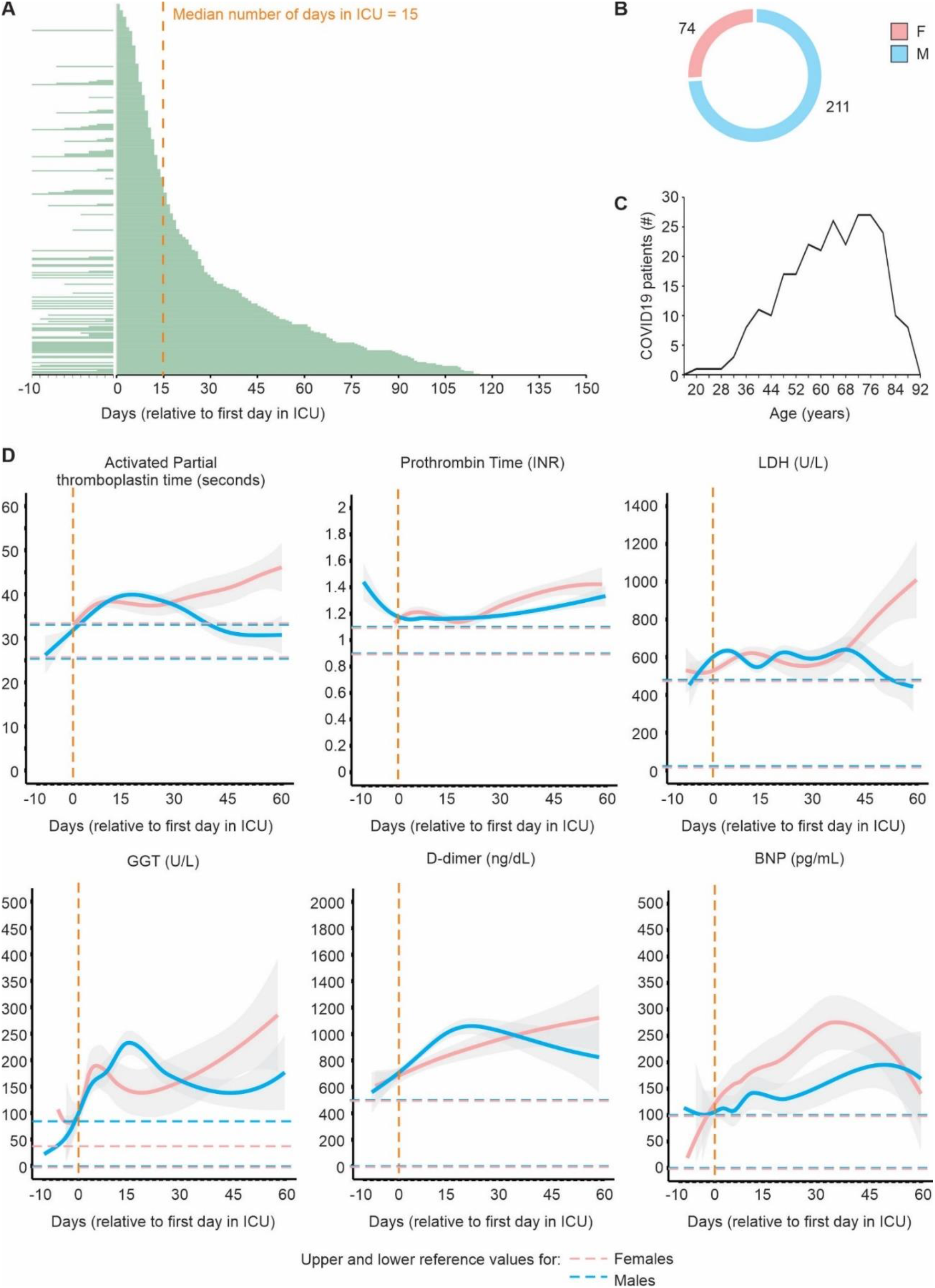
Time course analysis of COVID-19 patients in ICU. **A**. Number of days in ICU. Each patient is represented by a green line. The dashed vertical line shows the median number of days in ICU. **B**. Number of female and male patients in ICU. Males are represented in blue and females in pink. **C**. Age distribution of patients in ICU. **D**. Daily records of laboratory parameters in patients that went to ICU. Dashed vertical line represent the day of the first record in ICU. Values are shown in the y-axis and the units are displayed in parenthesis on top of each graph. The blue and pink lines represent the median values for males and females COVID-19 patients in CIU, respectively. The shaded area represents the 95% confidence interval. Horizontal dashed lines mark the upper and lower reference normal values for males (blue) and females (pink).

**Figure S2.**
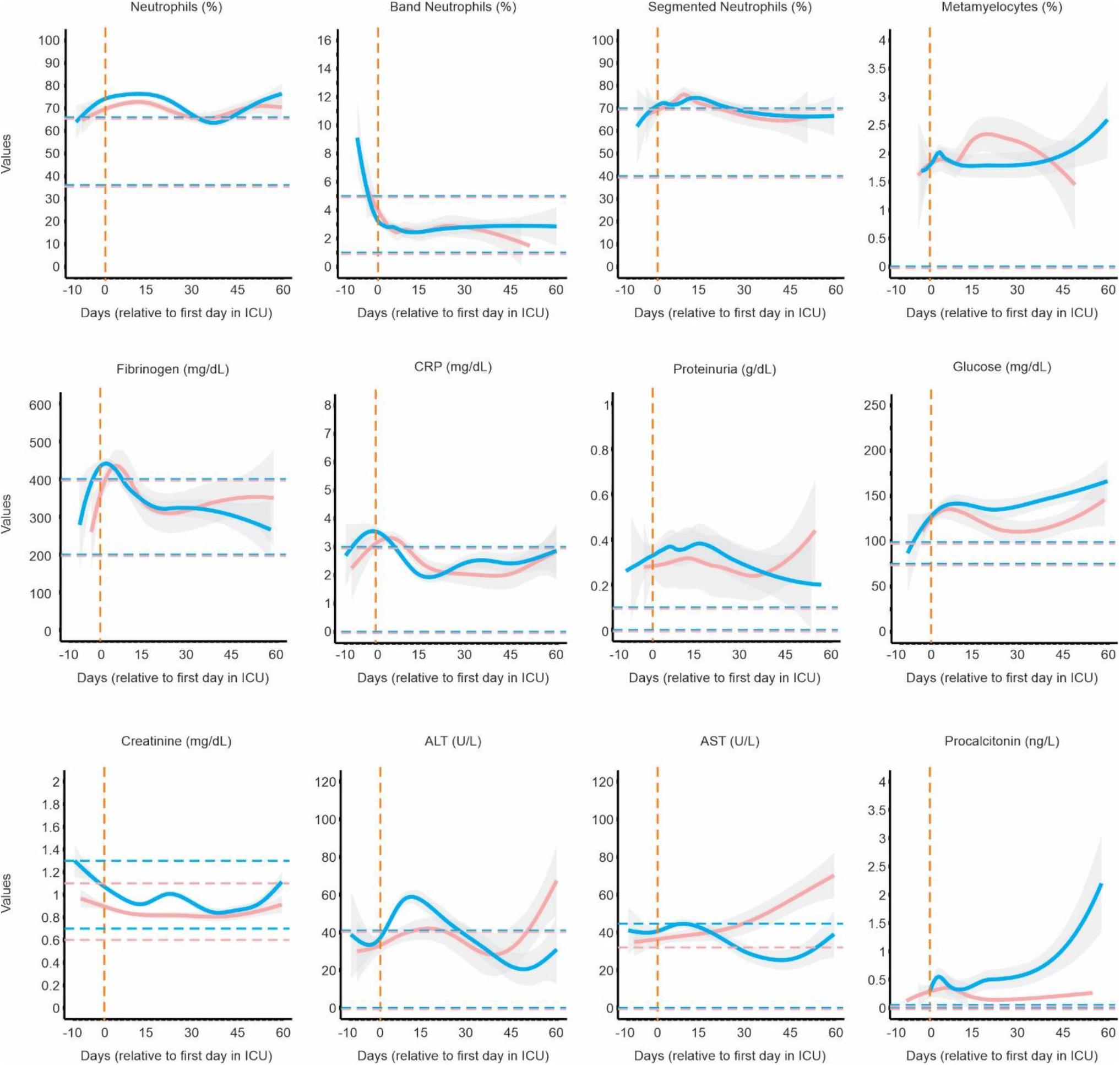
Time course analysis of COVID-19 patients in ICU, additional laboratory parameters. Daily records of laboratory parameters in patients that went to ICU. Dashed vertical line represent the day of the first record in ICU. Values are shown in the y-axis and the units are displayed in parenthesis on top of each graph. The blue and pink lines represent the median values for males and females COVID-19 patients in CIU, respectively. The shaded area represents the 95% confidence interval. Horizontal dashed lines mark the upper and lower reference normal values for males (blue) and females (pink).

Patients with COVID-19 may present with very different complications, such as liver or kidney impairment [7, 8], systemic inflammation [9] and coagulopathy [10]. We manually classified the laboratory parameters into biomarkers of disease activity for organ damage, problems in lipid or glucose metabolism, systemic inflammation, coagulation, acid-base balance in the blood gases, etc. (Table S2). Then, for each COVID-19 patient, we compared the earliest recorded levels of laboratory parameters after positive diagnosis with the respective reference values by sex. Alterations were similar between male and female COVID-19 patients that were admitted to ICU (Figure 5A). For instance, alterations in the frequency of immune cells were detected in 98% of male and female COVID-19 patients (Figure 5A). While 50.4% of male COVID-19 patients showed evidence of coagulopathy, only 39.7% of female patients had similar abnormalities (Figure 5A). When only ICU patients were considered, these numbers increased to 91.3% (males) and 82.8% (females) (Figure 5A). Furthermore, a large fraction of ICU patients showed evidence of elevated liver function tests and renal impairment, as well as disturbances of the acid-base balance, and systemic immune response (Figure 5A and 5B). Ten laboratory parameters were altered in a large fraction of ICU patients compared to all COVID-19 patients not admitted to the ICU. These included CRP, IL-6, neutrophils, ESR, fibrinogen, glucose, LDH, transferrin saturation (TS), albumin and total iron binding capacity (TIBC) (Figure 5C). These parameters further indicate that COVID-19 severity is indeed directly linked to disturbances in the coagulation and in the systemic immune response.

**Figure 5.**
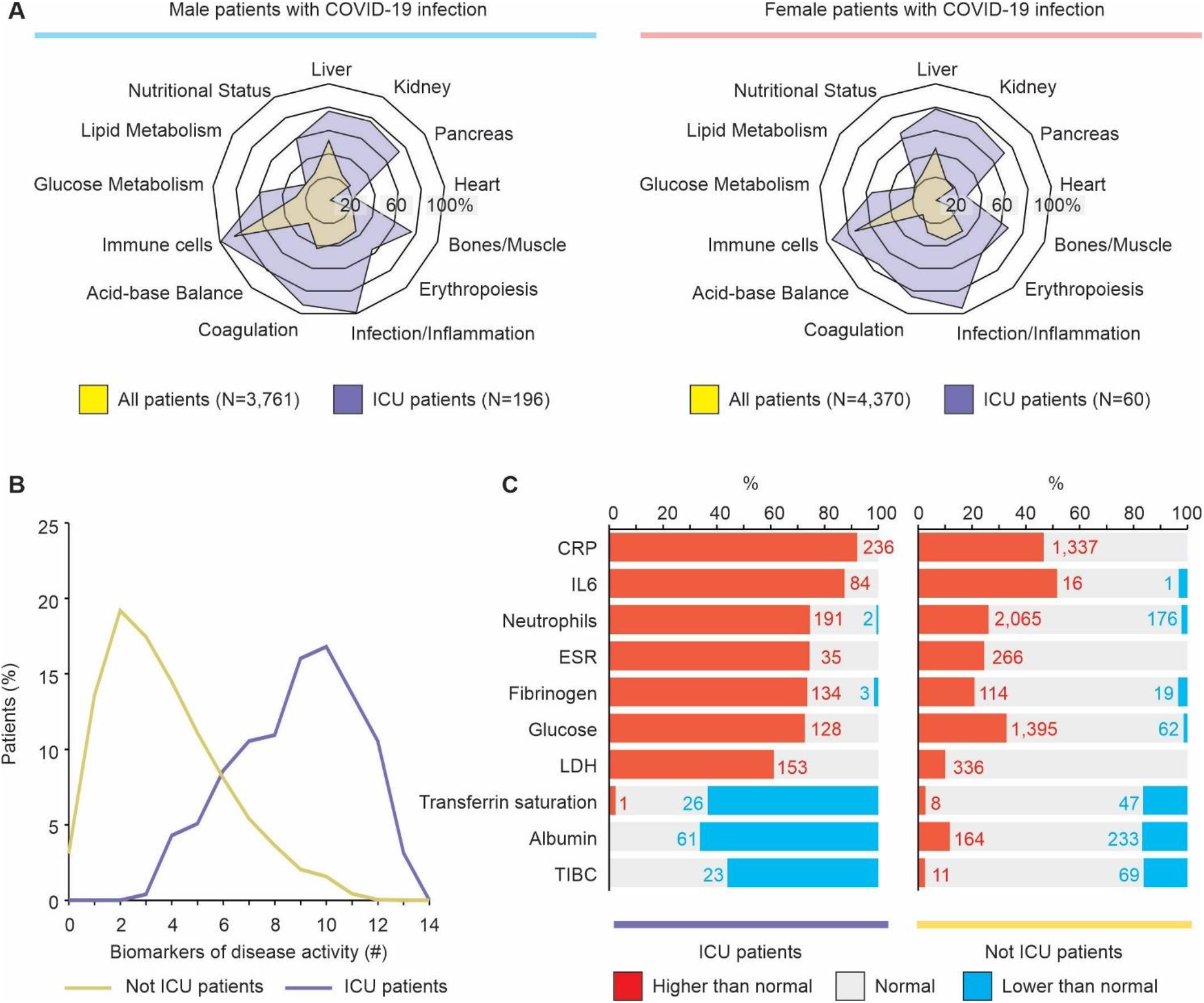
Biomarkers of disease activity of COVID-19 patients. **A**. Fraction of patients with altered biomarkers of disease activity. Alterations in at least one laboratory parameter of each biomarker of disease activity were detected for male (left) and female (right) COVID-19 patients that went (purple) or not (yellow) to ICU. The graph shows the fraction of subjects with alterations compared to the total number of subjects. **B**. Number of biomarkers of disease activity detected in COVID-19 patients. The purple and yellow lines indicate, respectively the fraction of COVID-19 patients that went or not to ICU. **C**. Laboratory parameters altered in ICU COVID-19 patients. Each bar represents the fraction of individuals that went (left) or not (right) to ICU which had values above (red) or below (blue) the normal levels for a specific parameter. Number of individuals with alterations are indicated.

## Discussion

Our in-depth analysis of routinely collected laboratory parameters in individuals with COVID-19 revealed that the changes seen are critically dependent on age and sex. This has significant implications for understanding the pathogenesis of COVID-19, as well as developing predictive models of SARS-CoV-2 infection and severe disease. For instance, we showed that elderly male patients have significantly more abnormal laboratory values, including higher inflammatory markers, compared to elderly females. The disproportionate male to female ratio of SARS-CoV-2 infection prevalence, morbidity and mortality has been reported around the world [11]. Severity and case fatality rates are significantly higher among men than women [12, 13]. Recent studies have considered the role of biological sex differences to explain the differences in outcome [14]. Sex hormones mediate differences in innate immune cells and functional responses to SARS, MERS, influenza and other viruses in the respiratory tract [15–17]. Our findings provide further support for exploring these hormonal influences in SARS-CoV-2 outcomes to explain why pre-menopausal women may be relatively protected from severe COVID-19 compared to men.

Increased levels of markers such as CRP, ferritin, fibrinogen, LDH and GGT could be detected in many COVID-19 patients. Ferritin and CRP have been widely described as consequences of the production of pro-inflammatory cytokines, such as IL-6, and the complement activation that may contribute to the systemic inflammatory response in patients with COVID-19 [18]. These changes were most marked in older men, although once patients were admitted to ICU, very similar profiles were observed between men and women.

Our findings from complete blood count analysis revealed that both male and female COVID-19 patients have low levels of platelets, basophils and eosinophils. Platelet numbers reduced with age in both men and women, with a similar reduction in COVID-19 patients compared to controls across age and sex. Thrombocytopenia is thought to be an early predictor of disease severity [19–21]. While lymphopenia is considered a feature of COVID-19, our analysis shows that the reduction is modest compared to patients without SARS-CoV-2 of a similar age and sex. Little is known about the roles of basophils and eosinophils in COVID-19. Eosinophils may enhance antiviral immunity during infection, and can activate viral antigen specific CD8+ T cells [22]. Also, previous studies suggest that female sex hormones regulate eosinophil numbers both in vitro and in vivo [15]. Using 989 patients, Li et al [23] reported that eosinopenia and elevated CRP effectively distinguish SARS-CoV-2-positive patients from SARS-CoV-2-negative patients with COVID-19-like symptoms. Using hematological data from over 30,000 people across all ages, we have demonstrated that eosinopenia and low levels of basophils are more accentuated in COVID-19 patients compared to other hematological parameters, especially in men older than 25 years of age.

The major limitation in our study is the lack of information regarding patient comorbidities and clinical outcome, other than admission to ICU. Patients admitted to ICU or that died from COVID-19, as well as those with comorbidities increasing the risk of severe disease [24], may display substantial variation in laboratory markers [25]. Overall, several abnormalities observed were accentuated in patients admitted to ICU, although we identified 10 markers that appeared to be more perturbed than others when compared to patients with COVID-19 not admitted to ICU. Interestingly, this included an elevated neutrophil count, which was not significantly different between COVID-19 and control patients overall. This may be related to increased bacterial co-infection in COVID-19 ICU patients, but also highlights potential differences between biomarkers associated with SARS-CoV-2 infection and those related to more severe disease. We also acknowledge that the differences seen in the temporal profiles may fall on a spectrum. Even though some laboratory values in COVID-19 patients fall in the normal range, they significantly differ from those patients without COVID-19.

We also did not have information regarding the cause of any illness in the control group, the individuals without SARS-CoV-2 infection. It is important to remember that the differences we describe in COVID-19 may be relative to individuals with other infections and not necessarily healthy controls. This is, however, also a key strength of our study when considering the utility of our findings for predictive models of SARS-CoV-2 infection and COVID-19 specific severity markers.

By analyzing both soluble and cellular markers in a large number of individuals, our study demonstrates that age and sex influence the results of the laboratory parameters measured and monitored in COVID-19 patients. Our findings should help guide both further studies into COVID-19 pathogenesis, as well as predictive models of SARS-CoV-2 infection and severe disease.

## Data Availability

All the original data is available at https://repositoriodatasharingfapesp.uspdigital.usp.br/.

## Data and materials availability

All the original data is available at https://repositoriodatasharingfapesp.uspdigital.usp.br/.

## Supplementary Materials

Tables S1 and S2

## References

1. Guan, W.J., et al., Clinical Characteristics of Coronavirus Disease 2019 in China. N Engl J Med, 2020.

2. Tay, M.Z., et al., The trinity of COVID-19: immunity, inflammation and intervention. Nat Rev Immunol, 2020. 20(6): p. 363–374.

3. Ng, S.C. and H. Tilg, COVID-19 and the gastrointestinal tract: more than meets the eye. Gut, 2020. 69(6): p. 973–974.

4. Feng, G., et al., COVID-19 and Liver Dysfunction: Current Insights and Emergent Therapeutic Strategies. J Clin Transl Hepatol, 2020. 8(1): p. 18–24.

5. Ronco, C., T. Reis, and F. Husain-Syed, Management of acute kidney injury in patients with COVID-19. Lancet Respir Med, 2020. 8(7): p. 738–742.

6. Zheng, Y.Y., et al., COVID-19 and the cardiovascular system. Nat Rev Cardiol, 2020. 17(5): p. 259–260.

7. Gholizadeh, P., et al., Alteration of Liver Biomarkers in Patients with SARS-CoV-2 (COVID-19). J Inflamm Res, 2020. 13: p. 285–292.

8. Hong, D., et al., Kidney manifestations of mild, moderate and severe coronavirus disease 2019: a retrospective cohort study. Clin Kidney J, 2020. 13(3): p. 340–346.

9. Gao, Y.M., et al., Cytokine storm syndrome in coronavirus disease 2019: A narrative review. J Intern Med, 2020.

10. Franchini, M., et al., COVID-19-associated coagulopathy. Diagnosis (Berl), 2020.

11. Scully, E.P., et al., Considering how biological sex impacts immune responses and COVID-19 outcomes. Nat Rev Immunol, 2020. 20(7): p. 442–447.

12. Qian, J., et al., Age-dependent gender differences of COVID-19 in mainland China: comparative study. Clin Infect Dis, 2020.

13. Michelozzi, P., et al., Mortality impacts of the coronavirus disease (COVID-19) outbreak by sex and age: rapid mortality surveillance system, Italy, 1 February to 18 April 2020. Euro Surveill, 2020. 25(19).

14. Ding, T., et al., Potential Influence of Menstrual Status and Sex Hormones on female SARS-CoV-2 Infection: A Cross-sectional Study from Multicentre in Wuhan, China. Clin Infect Dis, 2020.

15. Kadel, S. and S. Kovats, Sex Hormones Regulate Innate Immune Cells and Promote Sex Differences in Respiratory Virus Infection. Front Immunol, 2018. 9: p. 1653.

16. Karlberg, J., D.S. Chong, and W.Y. Lai, Do men have a higher case fatality rate of severe acute respiratory syndrome than women do? Am J Epidemiol, 2004. 159(3): p. 229-31.

17. Channappanavar, R., et al., Sex-Based Differences in Susceptibility to Severe Acute Respiratory Syndrome Coronavirus Infection. J Immunol, 2017. 198(10): p. 4046–4053.

18. Risitano, A.M., et al., Complement as a target in COVID-19? Nat Rev Immunol, 2020. 20(6): p. 343–344.

19. Zhou, F., et al., Clinical course and risk factors for mortality of adult inpatients with COVID-19 in Wuhan, China: a retrospective cohort study. Lancet, 2020.

20. Arachchillage, D.R.J. and M. Laffan, Abnormal coagulation parameters are associated with poor prognosis in patients with novel coronavirus pneumonia. J Thromb Haemost, 2020. 18(5): p. 1233–1234.

21. Bomhof, G., et al., COVID-19-associated immune thrombocytopenia. Br J Haematol, 2020. 190(2): p. e61–e64.

22. Samarasinghe, A.E., et al., Eosinophils Promote Antiviral Immunity in Mice Infected with Influenza A Virus. J Immunol, 2017. 198(8): p. 3214–3226.

23. Li, Q., et al., Eosinopenia and elevated C-reactive protein facilitate triage of COVID-19 patients in fever clinic: a retrospective case-control study. EClinicalMedicine, 2020: p. 100375.

24. Guan, W.J., et al., Comorbidity and its impact on 1590 patients with COVID-19 in China: a nationwide analysis. Eur Respir J, 2020. 55(5).

25. Velavan, T.P. and C.G. Meyer, Mild versus severe COVID-19: Laboratory markers. Int J Infect Dis, 2020. 95: p. 304–307.

